# Analytical Model of COVID-19 for lifting non-pharmaceutical interventions

**DOI:** 10.1101/2020.07.31.20166025

**Authors:** Garry Jacyna, James R. Thompson, Matt Koehler, David M. Slater

**Affiliations:** The MITRE Corporation: 7515 Colshire Drive, McLean, VA 22102

**Keywords:** COVID-19, Epidemiology, Mathematical model, Contact Network, Intervention

## Abstract

In the present work, we outline a set of coarse-grain analytical models that can be used by decision-makers to bound the potential impact of the COVID-19 pandemic on specific communities with known or estimated social contact structure and to assess the effects of various non-pharmaceutical interventions on slowing the progression of disease spread. This work provides a multi-dimensional view of the problem by examining steady-state and dynamic disease spread using a network-based approach. In addition, Bayesian-based estimation procedures are used to provide a realistic assessment of the severity of outbreaks based on estimates of the average and instantaneous basic reproduction number *R*_0_.

## 1. Introduction

Network structure plays an important role in disease spread. Most network models assume full mixing where all individuals are equally likely to become infected. However, most realworld networks have vertex degree distributions that are highly nonuniform. In the sections that follow, we derive a mathematical framework for determining both the steady-state and dynamic disease spread on complex networks using the concept of bond percolation. The analysis borrows liberally from the work of Pastor-Satorras and Vespignani [1] and Newman [2, 3, 4, 5].

Percolation theory is easily explained through analogy. Consider an old fashion coffee percolator consisting of two glass flasks, the lower flask for heating the water and the upper flask for holding the coffee grounds. As water is heated in the lower flask, it begins to ascend through the tube connected to the upper flask. Initially, the water penetrates only a small portion of the coffee grounds. Upon further heating, the water penetrates more of the coffee grounds. At some point, there is an abrupt transition where all of the grounds are saturated and convective mixing occurs. Percolation implies the existence of a long path connecting points separated by a distance on the order of the network size (in this case, the layer of coffee grounds). For a pandemic, this long path connects individuals together into a large cluster. It turns out that percolation is a critical phenomenon; that is, the onset of percolation occurs rapidly.

Most networks of sufficient complexity undergo phase transitions, where small components (outbreaks) suddenly coalesce into a giant component (pandemic) that extends across the entire network when one or more critical parameters, such as disease transmissibility are exceeded. Mean-field theory, a branch of statistical mechanics used to analyze physical systems with multiple components, can be used to characterize these regions [6]. The main idea is to replace all interactions on a component with an average or effective interaction. Insights into the behavior of a system can, therefore, be obtained at relatively low computational cost.

The flow chart in Fig. 1 outlines two analytical models suitable for describing the steady-state and time-dependent (dynamic) properties of disease progression on a social network. We present a third model that is used for estimating critical parameters (such as the basic reproduction number *R*_0_) from empirical outbreak case data. The steady-state model uses the theory of bond percolation to predict the outbreak size distribution prior to a pandemic, the size of the pandemic in terms of the proportion of affected individuals, and the risk of individual infection based on an individual’s contact network. We consider two types of non-pharmaceutical Interventions (NPIs)– uniform social distancing where a randomly selected portion of the population is sequestered and directed social distancing where individuals with the largest number of contacts are sequestered (which effectively targets super-spreaders). The dynamic model uses a degree-based approximation to the Susceptible, Infectious, or Recovered (SIR) model based on an approach outlined by Barthélémy and Pastor-Satorras [7]. We generalized the model to examine the effects of uniform social distancing, testing, and contact tracing on the proportion of susceptible individuals, infected individuals, outbreak cases, and the basic reproduction number (*R*_0_) as a function of time. In addition, assessments of various NPIs, testing, and contact tracing procedures can be determined in near real-time for both analytical and empirical networks derived from census data.

**Figure 1.**
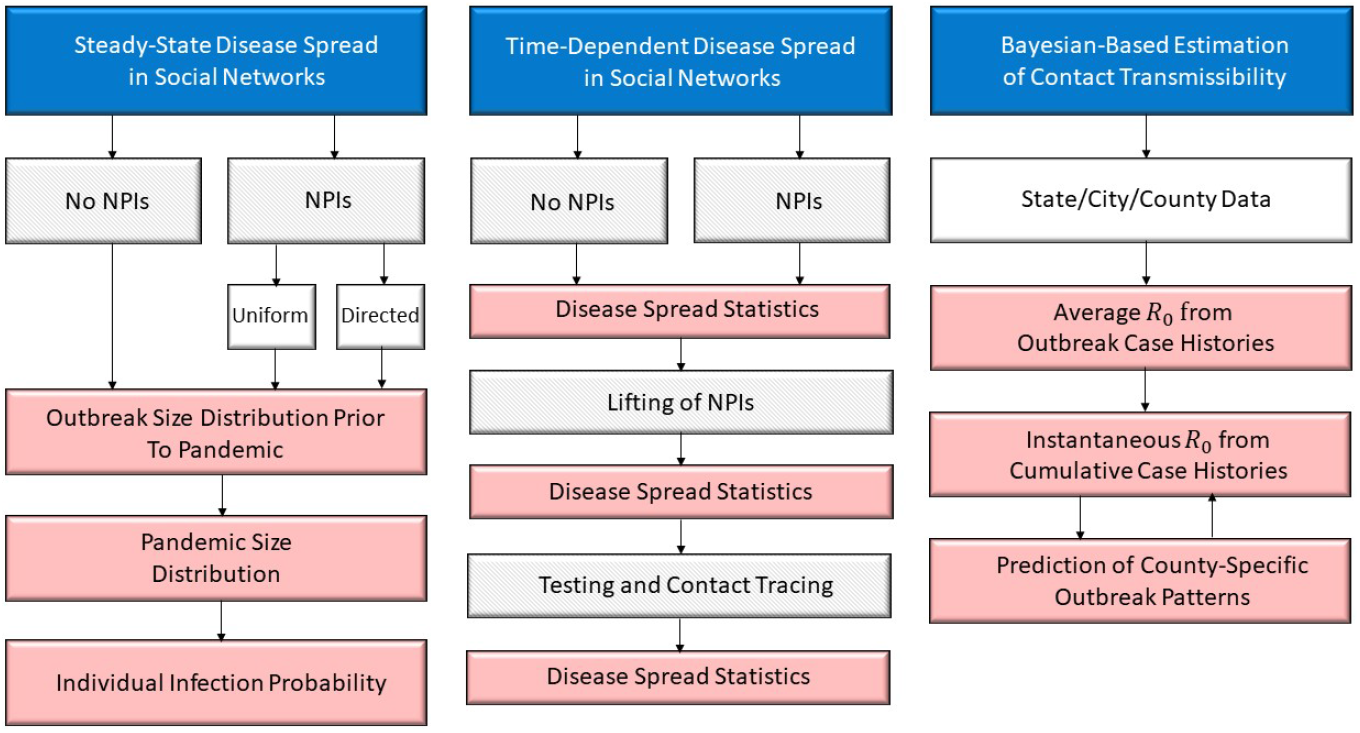
**Process Flow Diagram Detailing the Three Analytical Models Used in this Study**

The third model uses the degree-based SIR model and Bayesian estimation procedures to determine the average basic reproduction number (*R*_0_) from tabulated outbreak cases at a state, county, and city-wide level. Additionally, we use a particle-based filter to determine the instantaneous *R*_0_ over time. Together, these estimates can be used to predict the proportional number of infections and outbreak cases over time. The analytical models shown in Fig. 1 provide an overview of the mathematical framework derived in the present work. In the materials and methods section we first outline the steady-state disease spread on social contact networks as a function of the degree distribution of the community and the transmissibility of the disease. We then present the extension of the framework to capture the dynamic properties of disease spread and the implementation and lifting of NPIs. Next we introduce the Baysianbased estimation procedure for inferring network structure from empirical case data. In the results section we present examples for three different degree distributions and illustrate the different risk measures that can be derived from the mathematical framework. We also show how the Bayesian-based model can be used to predict cases and *R*_0_ for New York state using cases data collected by Johns Hopkins University [8]. We close with a brief discussion of the results and potential application of the framework.

## 2. Materials and methods

Throughout this section we use precise definitions of certain properties of epidemiological models based on Meyers et al., Bettencourt et al. and Chowell et al. [9, 10, 11].

- **Transmissibility** *T* is the average probability that an infectious individual will transmit the disease to a susceptible individual with whom they have contact
- **Critical Transmissibility** *T*_*c*_ is the minimum transmissibility required for an outbreak to become a pandemic. 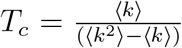 where ⟨*k*⟩ and ⟨*k*^2^⟩ are the mean and variance of the degree distribution of the contact network.
- **Basic Reproduction Number** *R*_0_ is the expected number of cases directly generated by one case in the population of susceptible individuals. It can be shown to be equal to the ratio of transmissibility to the critical transmissibility *R*_0_ = *T/T*_*c*_.
- **The Instantaneous Reproduction Number** *R*(*t*) = *R*_0_*s*(*t*)*/N* (*t*) where *s*(*t*) is the number of susceptible individuals at time *t* and *N* (*t*) is the total population.

Note that some studies refer to an effective reproductive number *R*_*e*_. In the definitions above, *R*_0_ depends on the degree distribution so there is no need to make this distinction. *R*_0_ and *R*_*e*_ can be thought of as interchangeable terms.

### 2.1. Steady-State Disease Spread in Social Networks

Here we outline the procedure for characterizing steady-state disease spread on complex networks using bond percolation. The analysis is based on the work of Pastor-Satorras and Vespignani [1] and Newman [2, 3, 4, 5]. Generalizations to the theory are made to include both uniform and directed social distancing.

Given the transmission rate *r*_*i,j*_ between node *i* and node *j* of a network graph and the infection time *τ*, the transmissibility is:

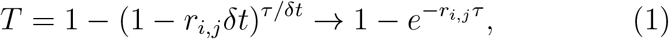

as *δt* → 0. Typically, *r*_*i,j*_ = *r*, where *r* and *τ* are independent random variables. The average transmissibility is then:

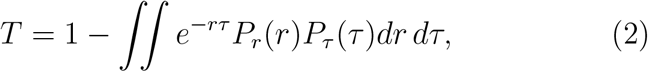

where *P*_*r*_(*r*) and *P*_*τ*_ (*τ*) are the respective probability density functions (pdfs). For simplicity, it is assumed that *P*_*r*_(*r*) = *δ*(*r* − *r*_0_) and *P*_*τ*_ (*τ*) = *δ*(*τ* − *τ*_0_) so that 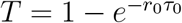.

For a randomly chosen vertex, let *p*_*k*_ denote the probability that this vertex has *k* edges. Define *G*_0_(*x*) as the generating function for the degree distribution of this vertex:

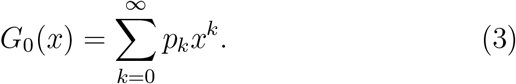

This function is similar to the characteristic function; that is, given a sum of *N* independent and identically-distributed (i. i. d.) random variables, the generating function is 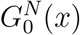.

Three types of social networks are examined. The ErdösRenyi network has a Poisson degree distribution of the form:

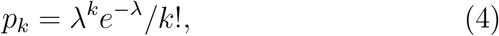

where *λ* is the mean. The exponential network is used as a proxy for an urban network and has a degree distribution of the form:

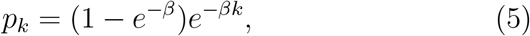

with parameter *β*. The power-law (Barabási-Albert) network has a majority of small degree links with a small minority of large degree links representing super-spreaders and a degree distribution of the form:

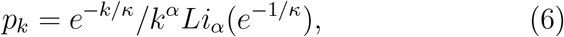

where *α* and *κ* are parameters and 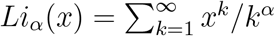 is the poly-logarithm function.

An important result by Feld is that the degree distribution of the first neighbor of a vertex is not the same as the degree distribution of vertices as a whole [12]. There is a greater chance that an edge will be connected to a vertex of high degree, in fact, in direct proportion to its degree. Let *q*_*k*_ denote the degree distribution of a vertex at the end of a randomly chosen edge. Then:

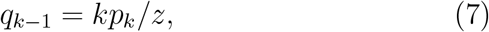

excluding the randomly-chosen edge, where *z* = Σ_*k*_ *kp*_*k*_. The corresponding generating function for this distribution is:

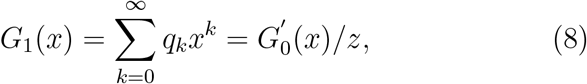

where 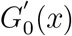 is the derivative of *G*_0_(*x*).

The transmissibility along each edge is taken into account by interpreting *T* in Eq. (2) as a probability (0 ≤ *T* ≤ 1). The probability that *m* out *k* edges is active is binomially distributed, so:

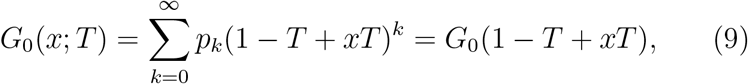

where, similarly, *G*_1_(*x*; *T*) = *G*_1_(1 − *T* + *xT*).

#### 2.1.1. Bond Percolation

Most complex networks experience phase transitions where small components suddenly coalesce into a giant component that extends across the entire network. The phase transitions of water as a function of temperature and pressure are examples. These regions can be described using mean-field theory and the generating functions *G*_0_(*x*) and *G*_1_(*x*). In order to apply mean field theory, it must be assumed that any finite component of connected vertices has no closed loops [5]. It can be shown that the probability of closed loops is on the order of 𝒪 (1*/n*), where *n* is the network size. As *n* → ∞, this means that all finite components have a tree-like (branching) structure.

Small outbreaks (percolation clusters) can be characterized as follows. Let *H*_1_(*x*) denote the generating function of the size distribution of the clusters at the end of the randomly chosen edge. Referring to the diagram in Fig. 2, the aggregate size of a cluster is the sum of all the clusters emanating from each vertex. This is a correct interpretation because there are no closed loops. For a vertex forming two clusters, the generating function of the size distribution is 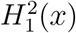, since two i. i. d. clusters are summed together.

**Figure 2.**
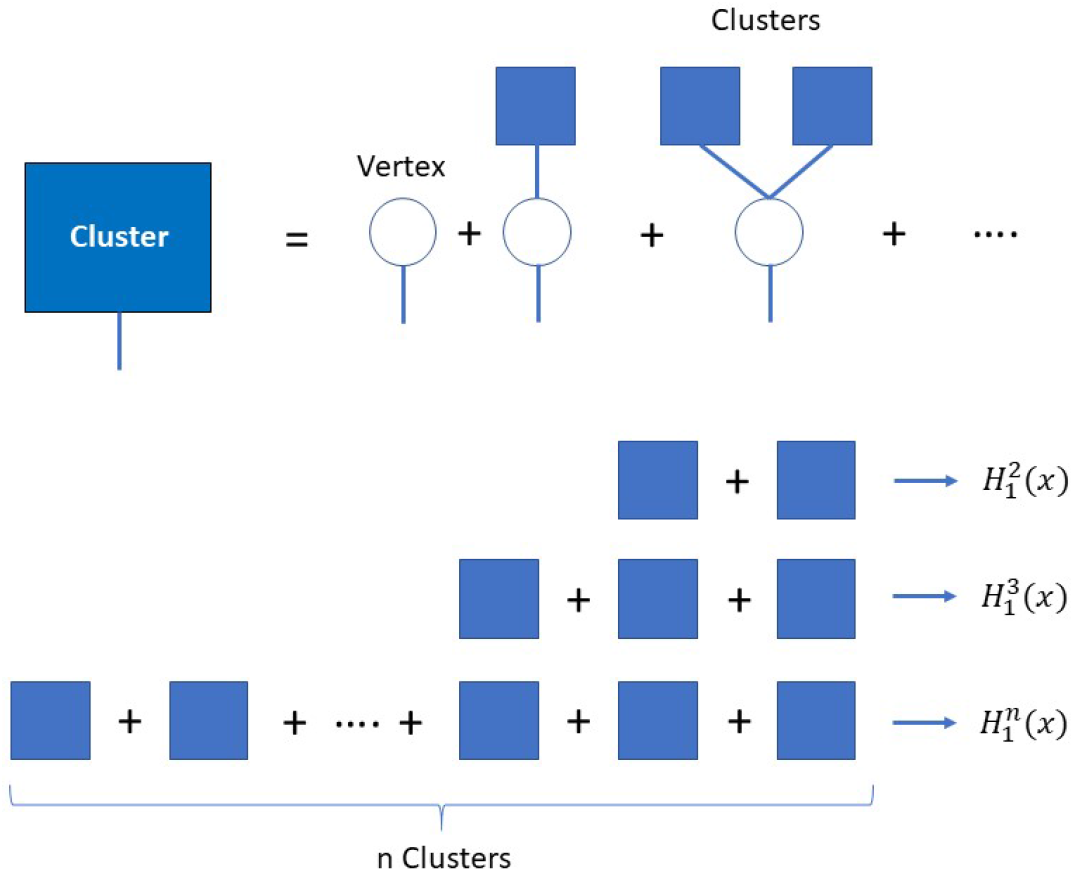
**Computing the size of percolation clusters using mean-field theory**

Similarly, for a vertex forming *n* clusters, the generating function of the size distribution is 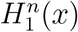. Therefore, in the limit of large network size:

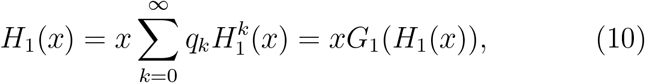

where *x* pre-multiplying the above expression is the originating vertex and *G*_1_(*x*) is as defined in Eq. (8) [3]. Similarly, defining *H*_0_(*x*) as the generating function of the size distribution of clusters for a randomly chosen vertex, then:

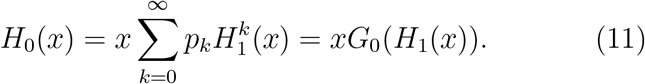

Modifying the above equations to include the transmissibility *T*:

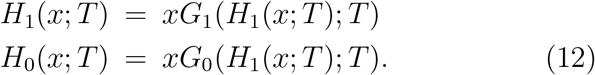

Equation (12) defines the bond percolation process and the analyses to follow.

#### 2.1.2. Small Outbreaks

The critical transmissibility *T*_*c*_ leading to a pandemic (giant component) can be determined by computing the average size of small outbreaks (small components not associated with the pandemic). Since 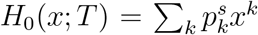, where 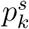 is the cluster size probability, the average size of the cluster is 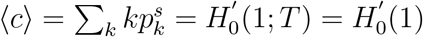. Newman shows that [2]:

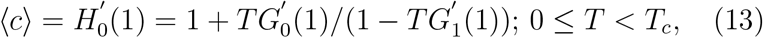

where *G*_0_(1) = *G*_1_(1) = 1. A phase transition occurs when ⟨*c*⟩ → ∞ or, from Eq. (13), when 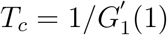 which implies:

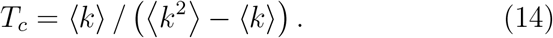

The basic reproduction number *R*_0_ can then be defined as *R*_0_ = *T/T*_*c*_, so *R*_0_ = 1 when *T* = *T*_*c*_.

In addition to the average outbreak size ⟨ *c* ⟩, the distribution of outbreak sizes can also be determined in a computationally efficient manner. Recalling that 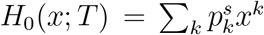 and letting *x* = *e*^2*πjm/M*^, the outbreak size distribution *p*_*k*_ is equivalent to the inverse discrete Fourier Transform (IDFT) of *H*_0_(*e*^2*pijm/M*^; *T*).

#### 2.1.3 Pandemic Size

The size of the pandemic above the critical threshold *T*_*c*_ can also be determined. However, the giant component spans the entire network, so there have to be closed loops. This invalidates the tree-like assumption that led to Eq. (12). As Newman points out, the problem can be approached indirectly [5]. Recall that 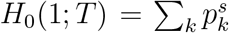 is the fraction of components *not* in the giant component. This implies that *P* (*T*) = 1 − *H*_0_(1; *T*) is the probability of a pandemic forming. Defining *v* = *H*_1_(1; *T*) and using Eq. (12):

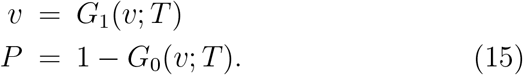

This is a fixed point problem. The average outbreak size not associated with the pandemic can also be determined. Repeating the derivation leading to Eq. (13) and noting that *H*_0_(1; *T*) = 1 − *P* (*T*), Newman shows that [3]:

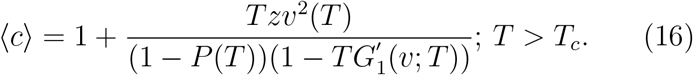

The risk of an individual infection is determined by noting that *v* in Eq. (15) is the probability that a node along a randomly chosen edge is not infected. For each edge, the probability of not getting infected is either *v* (contact not infected) or (1 − *T*)(1 − *v*) (contact is infected but does not transmit the infection). Thus, *p*(*T*) = *v* + (1 − *T*)(1 − *v*) = 1 − *T* − *vT*. If the individual has *k* contacts, then the risk of infection is [2]:

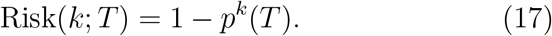

#### 2.1.4 Inclusion of NPIs

The known methodology outlined above is generalized to include social distancing. Specific applications including uniform and directed social distancing are then discussed. Let *b*_*k*_ denote the probability that a vertex of degree *k* is present. Define the generating function *F*_0_(*x*) as:

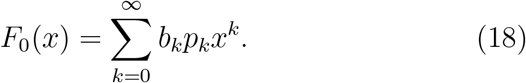

Similarly, 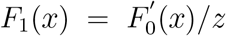. This is a generalization of the generating functions *G*_0_(*x*) and *G*_1_(*x*). Since *F*_0_(1) ≠ 1 and *F*_1_(1) ≠ 1, 1 − *F*_0_(1) is the probability that a randomly chosen vertex has no active edges. From Eq. (12), it follow that [3]:

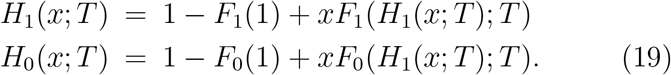

The average outbreak size prior to a pandemic with social distancing is similar to Eq. (13):

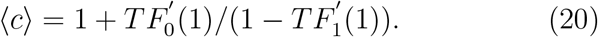

A phase transition leading to a pandemic occurs at the critical transmissibility 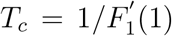, which follows directly from Eq. (20). Similarly, the outbreak size distribution follows directly from *H*_0_(*x*; *T*) by defining *x* = *e*^2*πjm/M*^ and using the inverse Fourier Transform.

The fraction of the population affected by the pandemic is similar to Eq. (15). Since *H*_0_(1; *T*) = 1 − *P* (*T*) and using Eq. (19):

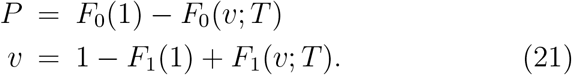

The average outbreak size not associated with the pandemic is analogous to Eq. (16) with *G*_0_(*x*) and *G*_1_(*x*) replaced by *F*_0_(*x*) and *F*_1_(*x*), respectively. Noting that *F*_0_(*v*; *T*) = *F*_0_(1) − *P*, 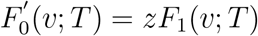, and *F*_1_(*v*; *T*) = *v* − 1 + *F*_1_(1):

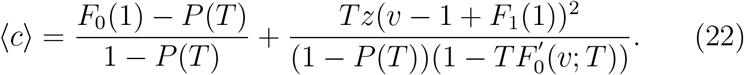

The risk of an infection during a pandemic is identical to Eq. (17) with *v* given by Eq. (21).

#### 2.1.5. Uniform Social Distancing

Let *b*_*k*_ = *b*; 0 ≤ *b* ≤ 1, so the probability that a vertex of degree *k* is active is *b*. Basically, social distancing is applied to every individual regardless of the number of contacts an individual may have. For this case, *F*_0_(*x*; *T*) = *bG*_0_(*x*; *T*) and *F*_1_(*x*; *T*) = *bG*_1_(*x*; *T*). The average outbreak size prior to a pandemic is identical to Eq. (13) with *T* replaced by *T*_eff_ = *bT*. This implies that the critical threshold with uniform social distancing is 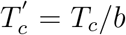. Since the basic reproduction number *R*_0_ = *T/T*_*c*_, the effective reproduction number is 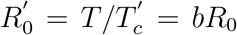. This means that the effective reproduction number 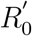 decreases with increased uniform social distancing. The outbreak size distribution similarly follows from Eq. (19).

The fraction of the population affected by the pandemic follows directly from Eq.(21). Letting *v*′ = (*v* −1+*b*)*/b* and noting that *G*_1_(*v*; *T*) = *G*_1_(1 − *bT* + *bTv*′):

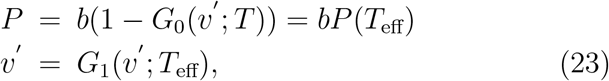

where *T*_eff_ = *bT*. This is identical to Eq. (15) with no social distancing except that the onset of the pandemic is shifted in accordance with *T*_eff_ and the affected population is reduced by a factor of *b*. Outbreaks not associated with the pandemic follow from Eq. (22). In particular:

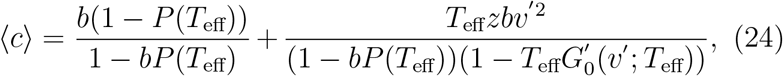

where *v*′ = (*v* − 1 + *b*)*/b*. Additionally, the risk of individual infection is given by Eq. (17) with 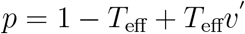.

#### 2.1.6. Directed Social Distancing

Here, it is assumed that:

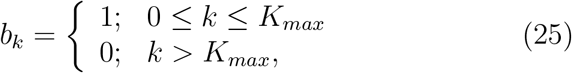

where all individuals are distanced with contact degree greater than *K*_*max*_. This implies that:

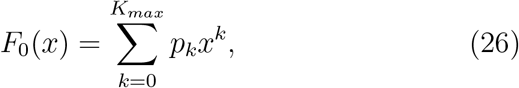

and 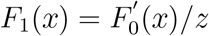. For a Poisson network:

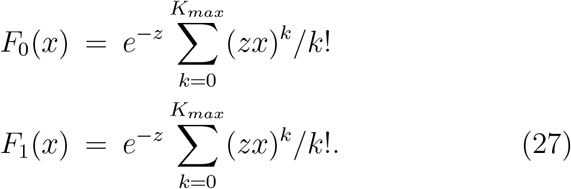

For an exponential network:

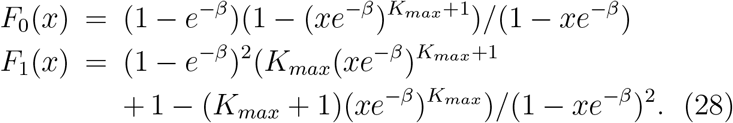

For a power-law network:

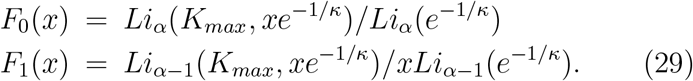

The average outbreak size prior to a pandemic is given by Eq. (20), where 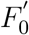 and 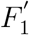 can be computed by taking respective derivatives of Eqs. (27), (28), and (29). Additionally, the fraction of the population affected by the pandemic is given by Eq. (21) together with Eqs. (27)-(29). The fractional number (*f*_*c*_) of nodes removed due to directed social distancing is related to *K*_*max*_ by:

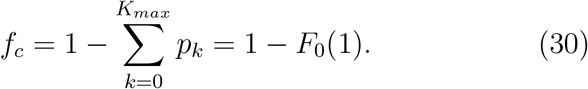

It is interesting to note that for the power-law network, if a small fraction of nodes *f*_*c*_ is removed, then the critical transmissibility 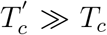, indicating a lack of phase transition or pandemic onset. This was discussed by Callaway and Newman in another context [4]. Both the average outbreak size not associated with the pandemic and the individual risk of infection are given by Eqs. (22) and (17), respectively.

#### 2.1.7. Calibration

The degree distribution *p*_*k*_ for each network is calibrated to have the same critical transmissibility *T*_*c*_. Since 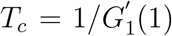 from Eq. (14), it follows that:

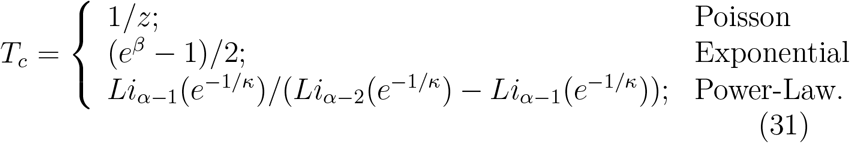

### 2.2. Time-dependent Disease Spread in Social Networks

In this section we expand the procedure to characterize time-dependent disease spread on complex networks using a stochastic SIR model. The analysis is based on the work of Barthélémy and Pastor-Satorras [7]. Generalizations to the theory are made to include social distancing, testing, and contact tracing.

#### 2.2.1. Introduction

A stochastic treatment of the time-dependent properties of epidemic models involves determining the probabilities for vertices to be in specific disease states. This is typically a difficult problem because it involves higher-order moments of probabilities that can only be approximated using moment-closure techniques, where moments are factored into pairwise moment products [5]. An alternative approach is degree-based approximation pioneered by Pastor-Satorras et al. This approximation assumes that all vertices of the same degree have the same probability of infection at any given time.

Consider the probability that vertex *A* becomes infected between times *t* and *t* + *dt*. To become infected, it must catch the disease from one of its neighbors, which requires that the neighbor be infected. The probability of a neighbor being infected is *x*_*k*_, where *k* is the excess degree of the neighbor (recall that the excess degree distribution is *q*_*k*_). So, the average probability of a neighbor being infected is 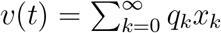. The total probability of transmission from a single neighbor is *βv*(*t*)*dt*, where *β* is the contact rate. The probability of transmission from any neighbor is *βkv*(*t*)*dt*, where *k* is the number of neighbors associated with vertex *A*. Thus, the rate of change in the probability that a vertex with degree *k* is susceptible (*s*_*k*_) is simply:

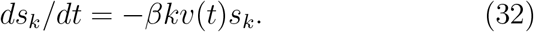

Similarly, the probability that a vertex with degree *k* is infected (*x*_*k*_) is:

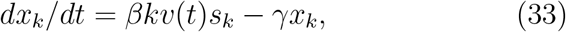

where *γ* = 1*/T*_*r*_ is the recovery rate. Finally, the probability that a vertex of degree *k* has recovered (*r*_*k*_) is:

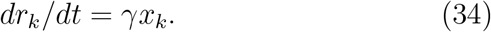

A general solution to Eqs. (32)-(34) is of the form [5]:

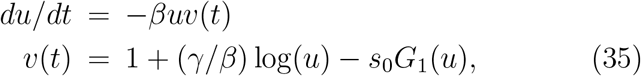

where *s*_0_ = *s*_*k*_(0). Given *u*(0) = 1, Eq. (35) can be numerically solved for *u*(*t*) and the average probability of infection *v*(*t*) can be determined. In addition, the average susceptibility probability *s*(*t*) can be computed from the degree distribution of a randomly chosen vertex:

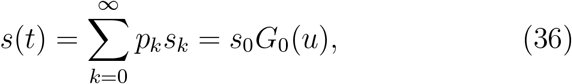

where *G*_0_(*x*) is the moment generating function for *p*_*k*_.

The function *u*(*t*) in Eq. (35) has a particularly interesting interpretation. A fixed point occurs when *du/dt* = 0 or when 1 + (*γ/β*) log(*u*) − *s*_0_*G*_1_(*u*) = 0. If it is assumed that *u* ≈ 1, then:

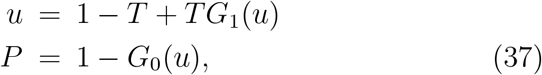

where the pandemic size *P* = *w*(∞) and 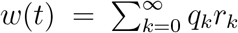 is the average recovery probability. Assuming *β/γ* ≪ 1, the transmissibility is approximately *T* ≈ *β/γ*. If *u* = 1 − *T* + *Tv*, then Eq. (15) and Eq. (37) are identical under the above assumptions. Therefore, the dynamic SIR model is consistent with bond percolation results at steady state.

#### 2.2.2. Uniform Social Distancing

It is assumed that no NPIs are active over a period 0 ≤ *t* ≤ *t*_1_. Over a period *t*_1_ *< t* ≤ *t*_2_, uniform social distancing is implemented where a fraction (1 − *b*) of the population is sequestered. Over the period *t*_2_ *< t* ≤ *t*_3_, the NPI is lifted. The first period corresponds to the analysis outlined in the previous section.

For the second period that includes social distancing, the stochastic SIR equations for the fraction of the population (*b*) not sequestered are:

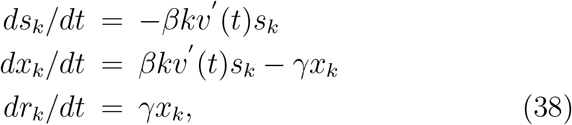

where 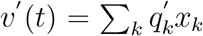 and 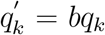. Letting 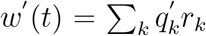, it follows that *dw*′ */dt* = *γv*′ (*t*). Substituting this expression into Eq. (38) and simplifying:

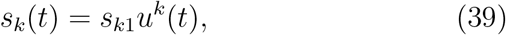

where 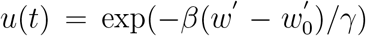and *s*_*k*1_ are the fractional number of susceptible individuals with *k* contacts remaining at the end of the first period (*t* = *t*_1_). Since *x*_*k*_ = 1 − *s*_*k*_ − *r*_*k*_, 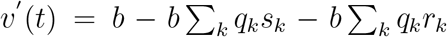. Now, 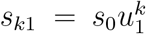, where *u*_1_ = *u*(*t*_1_), so 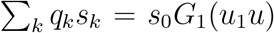. Also, 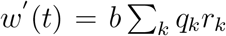. Using the expression for 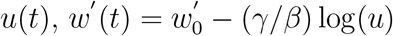, where 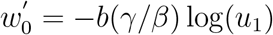. Combining results:

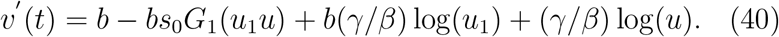

Finally, since *dw*′ */dt* = −(*γ/βu*)*du/dt* and *dw*′ */dt* = *γv*′ (*t*):

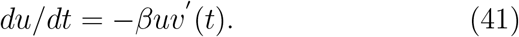

The total average infection probability *v*_*T*_ (*t*) needs to include the sequestered fraction (1 − *b*) of the population. However, the average infection probability among the sequestered population is 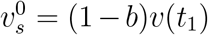, where *v*(*t*_1_) is determined from Eq. (35). In particular:

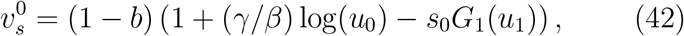

where *u*_1_ = *u*(*t*_1_). These infected individuals recover at a rate *γ* since, by definition, they are not in contact with each other. So:

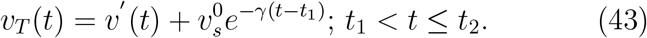

Note that at *t* = *t*_1_, *v*_*T*_ (*t*_1_) = *v*(*t*_1_), so continuity is maintained. Finally, the fractional number of susceptibles during the NPI period is the sum of the sequestered and non-sequestered fraction of the population:

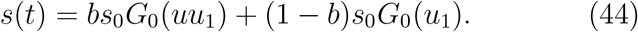

For the third period when social distancing is lifted, the stochastic SIR equations are identical to Eqs. (32)-(34) except that the initial conditions are different. Let *n*_*k*_ denote the fraction of the population with *k* contacts remaining. However, *n*_*k*_ = *b* + (1 − *b*)*s*_*k*1_, i.e., the original non-sequestered fraction (*b*) and the sequestered fraction ((1 − *b*)*s*_*k*1_), where 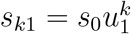.

Let 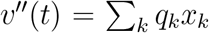 denote the average infection probability, where *x*_*k*_ = *n*_*k*_ − *s*_*k*_ − *r*_*k*_. Now, *s*_*k*_ has the form:

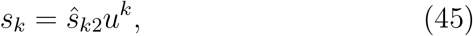

Where 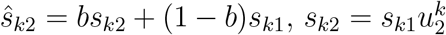, and *u*_2_ = *u*(*t*_2_). The various summations entering into the calculation of *v″* (*t*) are of the form:

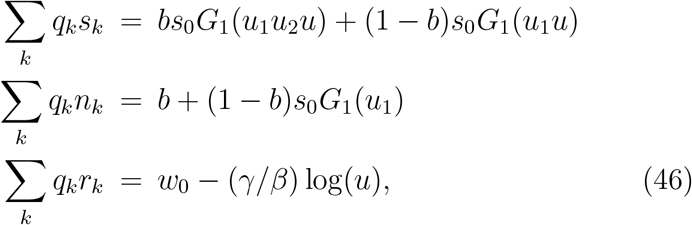

where *w*_0_ = −(*γ/β*) log(*u*_2_)−*b*(*γ/β*) log(*u*_1_). Combining results:

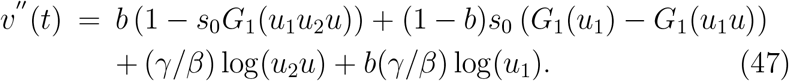

Note that *v″* (*t*_2_) = *v*′ (*t*_2_), as required. The fractional number of susceptibles is given by:

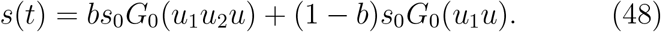

### 2.2.3. Testing and Contact Tracing

For periods *t > t*_3_, a fairly simple model is used to capture the relevant details of delayed testing and contact tracing based on a generalization of work by Young and Ruschel [13]. The modified stochastic SIR model is:

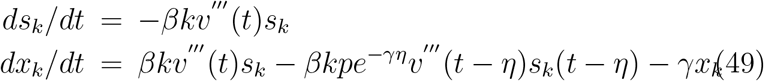

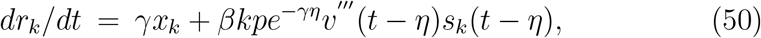

where *η* is the delay in testing after becoming infectious and, as before, 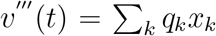. Rather than assuming that infectious individuals are tested, tracked and sequestered immediately with probability *p*, there is a delay *η* in their sequestration. Recovered individuals are not added back into the pool of susceptible individuals since it is assumed that they have developed immunity to the disease.

A general solution to Eq. (50) can be determined as follows. Integrating the first expression produces 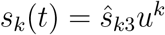, where:

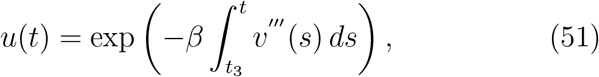

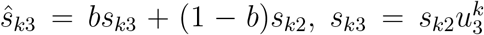, and *u*_3_ = *u*(*t*_3_). From Eq. (51), it follows that:

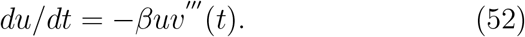

Averaging the second expression in Eq. (50):

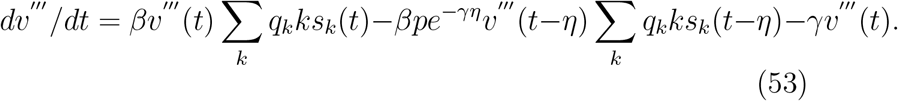

Noting that 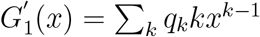:

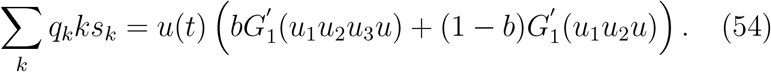

Therefore, Eq. (53) can be rewritten as:

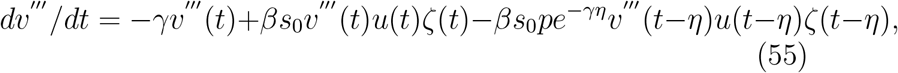

where 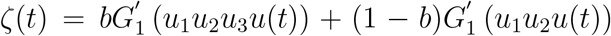. Equations (52) and (55) describe a set of consistent equations for numerically computing *v*‴ (*t*) and *s*(*t*), where:

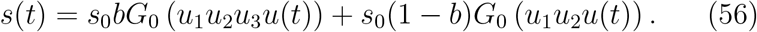

Note that Eq. (55) is initialized by setting *v*‴ (*t*_3_) = *v″* (*t*_3_).

### 2.3. Bayesian-based Estimation of Contact Transmissibility

We now outline methods for estimating the contact transmissibility from outbreak case data using a stochastic SIR model and Bayesian estimation methods. The first approach estimates the average transmissibility *T* or basic reproduction number *R*_0_ using either analytical or empirical networks where the degree distribution is known or estimated. The second approach estimates the instantaneous transmissibility *T* (*t*) or basic reproduction number *R*_0_(*t*) using nonlinear tracking methods based on particle-based filtering where explicit network structure is not required.

#### 2.3.1. Average Transmissibility

The probability that a vertex of degree *k* will experience an outbreak is the probability that it gets infected and then recovers. From Eq. (33):

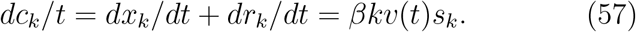

The average outbreak case probability *C*(*t*) is determined by averaging Eq. (57):

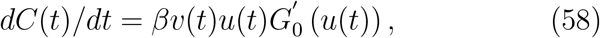

where *u*(*t*) is given by Eq. (35) and 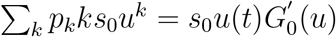.

Define Δ*C*(*t* + *ξ*) = *C*(*t* + *ξ*) − *C*(*t*) as the fractional change in outbreak cases in the interval [*t, t* + *ξ*]. Then, from Eq. (58):

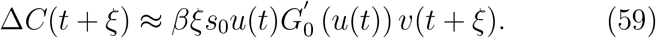

Now, returning to Eq. (33), averaging, and integrating the results from *t* to *t* + *ξ*, assuming that *ξ* ≪ 1:

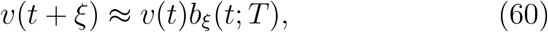

where 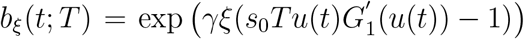 and *T* = *β/γ*. Finally, substituting this expression into Eq. (59):

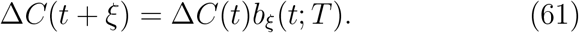

This implies a linear relationship between the fractional change in outbreak cases between time steps *ξ*.

The change in the number of outbreak cases Δ*N* (*t*) is Δ*N* (*t*) = *NC*(*t*), where *N* is the population size. Assume that Δ*N* (*t*) is integer-valued and let the measurements comprise the collection {Δ*N*_1_, Δ*N*_2_,…, Δ*N*_*J*_}, where Δ*N*_*i*_ = Δ*N* (*t*_*i*_). The resulting likelihood function can be factored as:

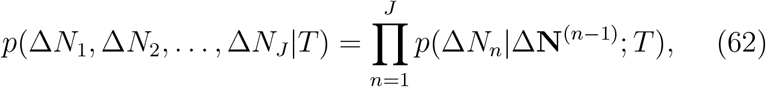

where Δ**N**^*n*^ = [Δ*N*_1_, Δ*N*_2_,…, Δ*N*_*n*_]. This equation can be simplified by noting that Δ*N*_*i*+1_ = *b*_*ξ*_(*t*_*i*_; *T*)Δ*N*_*i*_, so Δ*N*_*i*_ defines a Markov process. Therefore:

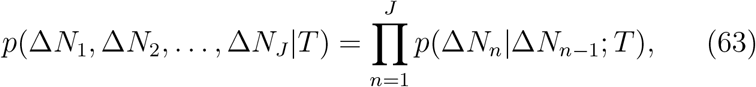

where *p*(Δ*N*_1_|Δ*N*_0_; *T*) = *p*(Δ*N*_1_).

In general, *p*(Δ*N*_*n*_|Δ*N*_*n*−1_; *T*) is not known. Bettencourt points out that the maximum entropy density is the preferred density when only the mean is known [10]. This turns out to be the Poisson density:

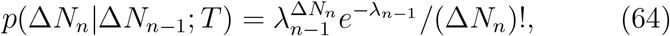

where *λ*_*n*_ = *b*_*ξ*_(*t*_*n*_; *T*)Δ*N*_*n*_. Taking the logarithm of Eq. (63), using Eq. (64), and eliminating terms independent of *T*, − log *p*(Δ*N*_*n*_|Δ*N*_*n*−1_; *T*) is proportional to:

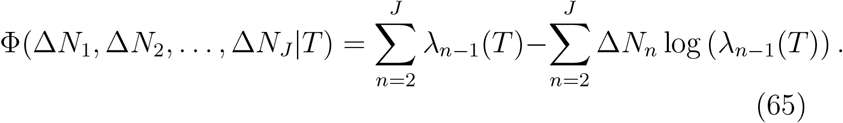

A maximum likelihood estimate of the transmissibility *T* or basic reproduction number *R*_0_ is equivalent to finding a *T* such that Φ(Δ*N*_1_, Δ*N*_2_,…, Δ*N*_*J*_ |*T*) is minimized.

Although a Poisson density is used in deriving the conditional likelihood function in Eq. (64), it has been observed that the number of differential outbreak cases Δ*N* (*t*_*i*_) is overdispersive, i.e., the variances are larger than expected. It can be shown that Consul’s generalized Poisson distribution is a better match to the data. This density has the form [14]:

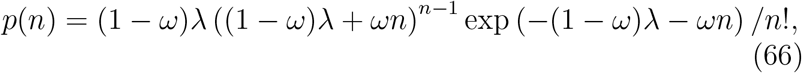

where 0 ≤ *ω <* 1 is the dispersion parameter and *λ* is the mean. Euler’s difference formula can be used to show that *p*(*n*) is a valid density (∑_*n*_ *p*(*n*) = 1).

#### 2.3.2. Estimating Instantaneous Transmissibility

Define the instantaneous basic reproduction number *R*_0_(*t*) as 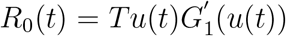, which follows from Eq. (60), so that *b*_*ξ*_(*t*; *T*) = exp (*γξ*(*R*(*t*) − 1)). This equation has the advantage of being network independent. Now, consider the vector of estimates: **R**^*J*^ = [*R*_(_*t*_1_), *R*(*t*_2_),…, *R*(*t*_*J*_)] and the vector of measurements Δ**N**^*J*^, where Δ**N**^*J*^ = [Δ*N*_1_, Δ*N*_2_,…, Δ*N*_*J*_]. The goal is to estimate the posterior density *p*(**R**^*J*^ |Δ**N**^*J*^). Because the likelihood function is a nonlinear function of *R*_0_, a linear Kalman filter cannot be used. In fact, an extended Kalman filter is not robust enough to handle the rapid fluctuations in the differential outbreak case histories. Therefore, a Bayesian approach is used where the posterior probability density of the state is constructed from the data. However, this density may be difficult to evaluate using kernel or grid-based estimation procedures. Therefore, the density is approximated using sampling procedures.

In order to illustrate the procedure, the following simple problem is considered. Suppose one is required to evaluate the N^*th*^ moment of *p*(*x*|*z*):

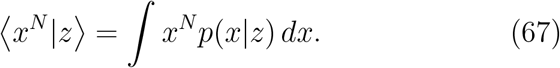

Assume a proposal density *q*(*x*|*z*) that is relatively easy to sample from. These samples are denoted by *x*^*i*^ ∼ *q*(*x*|*z*) such that 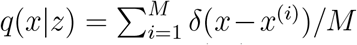, where *δ*(*x*) is the Dirac delta function. Equation (67) can be rewritten as:

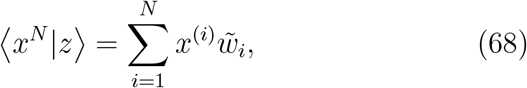

where 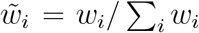 and *w*_*i*_ = *p*(*x*^(*i*)^ | *z*)*/q*(*x*^(*i*)^ | *z*). Therefore, the N^*th*^ moment can be approximated by weighting samples *x*^(*i*)^ from a proposal density *q*(*x*|*z*) by a set of importance weights 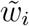.

The same procedure can be used to estimate the posterior density. In this case *w*_*i*_ = *p*(**R**^*J*(*i*)^ | ΔN^*J*^)*/q*(**R**^*J*(*i*)^ |ΔN^*J*^) for a suitably chosen proposal density *q*(**R**^*J*^ |Δ**N**^*J*^). Now, using Bayes theorem:

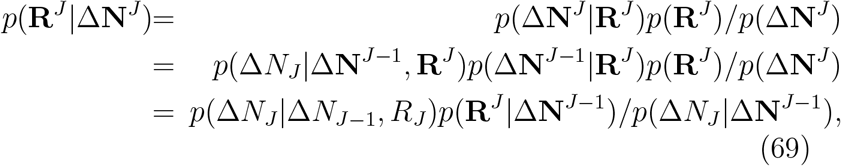

where it is assumed that *p*(Δ*N*_*J*_ |Δ**N**^*J*−1^, **R**^*J*^) = *p*(Δ*N*_*J*_ |Δ*N*_*J*−1_, *R*_*J*_). In addition, assume that *R*_*J*_ is Markov so that *p*(**R**^*J*^ |Δ**N**^*J*−1^) = *p*(*R*_*J*_ |*R*_*J*−1_)*p*(**R**^*J*−1^|Δ**N**^*J*−1^). From Eq. (69):

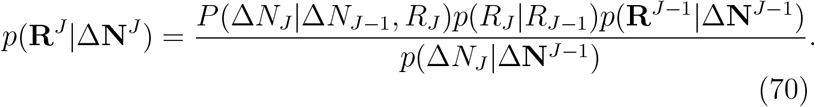

The proposal density can be factored into the following form 504 using product of conditional densities:

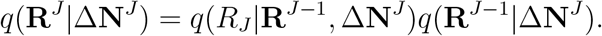

Suppose that *q*(*R*_*J*_ |**R**^*J*−1^, Δ**N**^*J*^) = *q*(*R*_*J*_ |*R*_*J*−1_) and *q*(**R**^*J*−1^|Δ**N**^*J*^) = *q*(**R**^*J*−1^|Δ**N**^*J*−1^). Using the definition of *w*_*i*_ above:

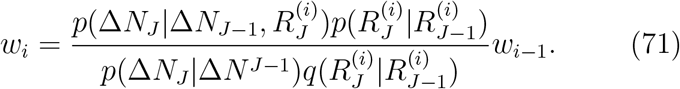

For a simple particle filter examined here, 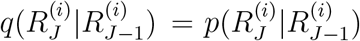, so that:

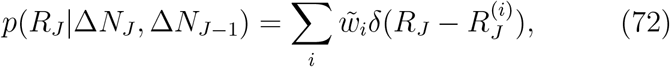

where 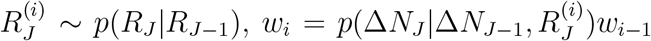, and 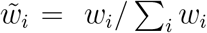. In order to prevent particle degeneracy (collapse to a few particles), the nonuniform measure in Eq. (72) is replaced by a uniform measure by computing the cumulative distribution function (cdf) of 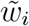 and uniformly sampling to produce 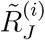 such that 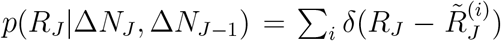 [15].

The state transition (sampling) density *p*(*R*_*J*_ |*R*_*J*−1_) can be determined as follows. Augment the state *R*_*J*_ such that 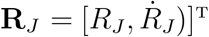 and assume a linear projection of the form:

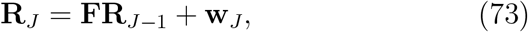

where 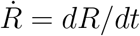 and:

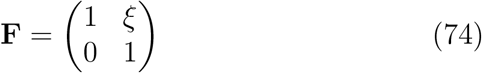

and **w**_*J*_ is a zero-mean Gaussian noise vector. Assume that 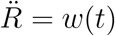, where *w*(*t*) is a zero-mean white Gaussian noise process and 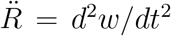. This constitutes a random “acceleration” model. It can be shown that the corresponding covariance matrix associated with the state **R**_*J*_ in Eq. (73) is:

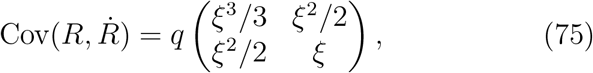

where *q* is a variance-like term. It then follows that *p*(*R*_*J*_ |*R*_*J*−1_) is a zero-mean Gaussian density with covariance Cov 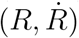. The likelihood function is computed from Eq. (64).

## 3. Results and Discussion

### 3.1. Steady-State Disease Spread

Some illustrative results using the steady-state disease spread model are described below for three types of contact networks: Poisson, Exponential, and Power-law. For these examples, the critical transmissibility is *T*_*c*_ = 0.049. This means that if the observed transmissibility *T* = *T*_*c*_, then *R*_0_ = 1, i.e., *T*_*c*_ is the critical value above which the pandemic is self-sustaining. For comparative purposes, each contact network has the same critical threshold *T*_*c*_.

Fig. 3 depicts the outbreak distribution size prior to a pandemic when *R*_0_ = 0.8 with no NPIs. This is the predicted number of people infected by a small outbreak. Both the Poisson and exponential (urban) contact networks have similar size distributions, where large outbreak sizes (*>* 20) are unlikely. The power-law contact network is highly peaked for small outbreak sizes as expected. Although diminishingly small for larger outbreak sizes, it is not zero because there are a minority of super-spreaders with large contact degree. Fig. 4 depicts the fraction of the population affected by a pandemic as a function of the basic reproduction number *R*_0_ when *R*_0_ *>* 1 with no NPIs. Communities have diverse experiences based on their contact patterns. The Poisson contact network has the greatest fraction of the population affected by a pandemic because individuals in a group are equally likely to become infected and to infect others. For an exponential (urban) contact network, there is a 50% reduction in the number of infected individuals compared to a well-mixed population (40% versus 80%) for *R*_0_ = 2. For a power-law network, only 5% of the population is affected by the pandemic for *R*_0_ = 2. Therefore, outbreaks are consistently less likely to reach pandemic proportions.

**Figure 3.**
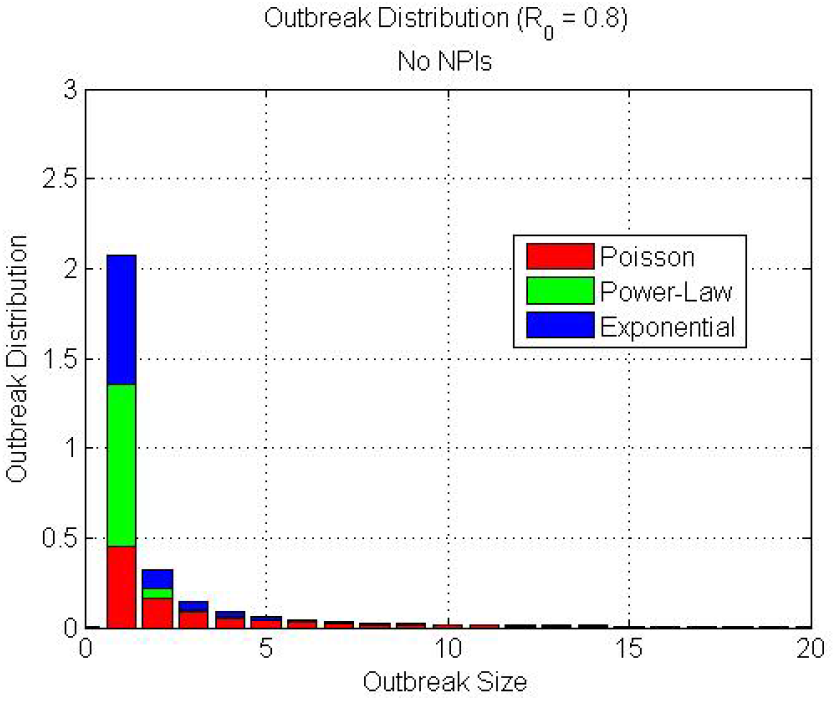
**Outbreak size distribution prior to a pandemic (***R*_0_ = 0.8**) for three types of contact networks**

**Figure 4.**
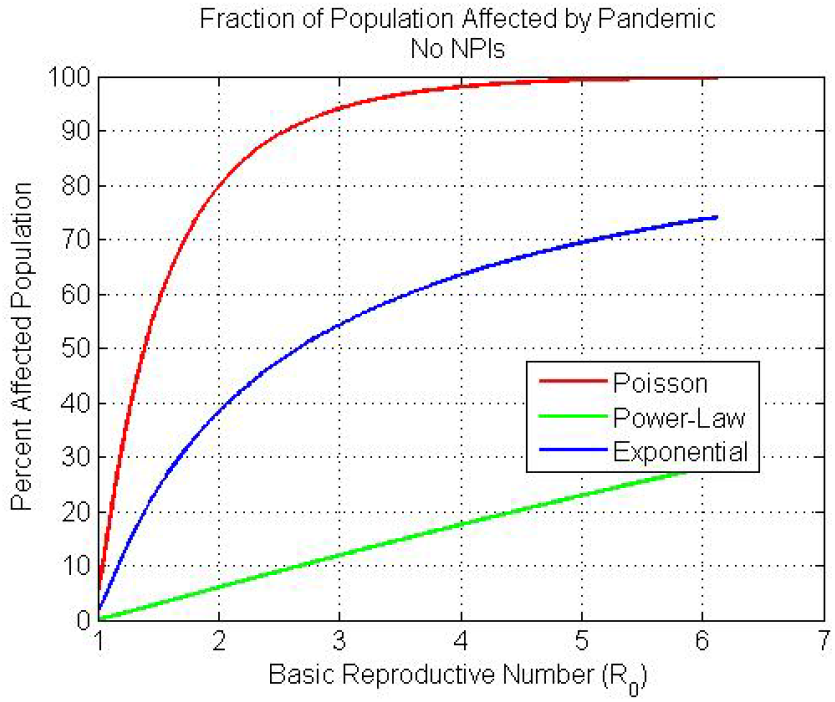
**Pandemic size (or fraction of individuals affected) as a function of** *R*_0_ **for three types of contact networks**

There are outbreaks that can occur outside the main pandemic cluster, although they are relatively small. This is not predicted from SIR or Susceptible, Exposed, Infected, or Recovered (SEIR) models. Fig. 5 shows the average number of people infected by outbreaks outside the main pandemic cluster. This average outbreak size can be determined as a function of *R*_0_ for the three contact networks. For *R*_0_ *>* 2 all three contact networks have low average outbreak sizes (less than 2) outside the main pandemic cluster. As expected, there are more outbreaks near *R*_0_ = 1 because less of the population is affected by the main pandemic and there is more opportunity for infections to spread outside the main pandemic cluster. Note that the power-law network has a larger outbreak size for larger *R*_0_ because there are a minority of super-spreaders. Fig. 6 illustrates the individual risk of infection when *R*_0_ = 2 based on the number of social contacts. The Poisson contact network shows the most risk of individual infection. For ten contacts, the risk of infection is approximately 55%. The exponential (urban) network shows an individual infection risk of approximately 45% for ten contacts. The power-law contact network shows the smallest individual risk of infection, roughly 20% for ten contacts.

**Figure 5.**
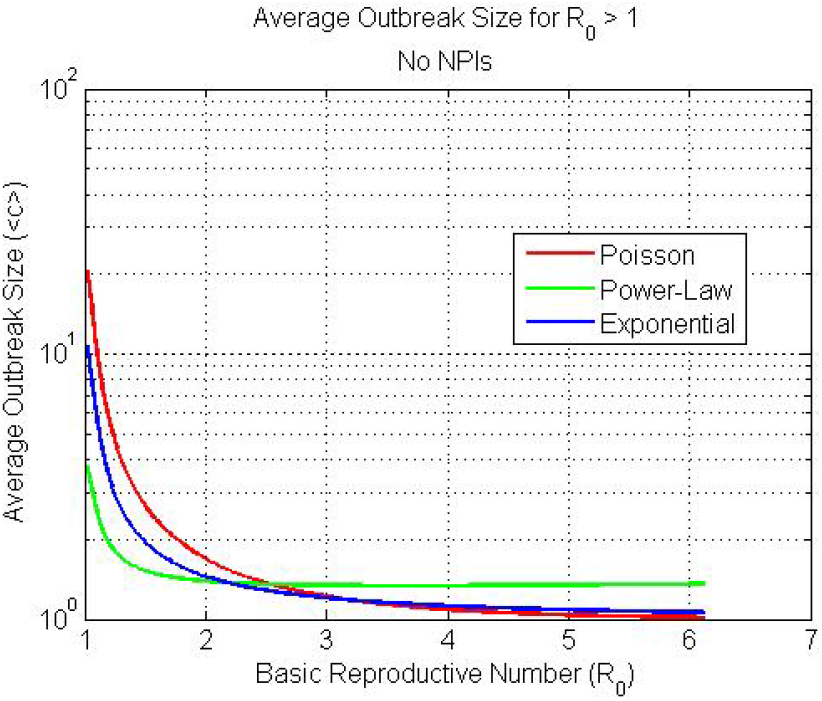
**Average outbreak size removed from the main pandemic cluster for three types of contact networks**

**Figure 6.**
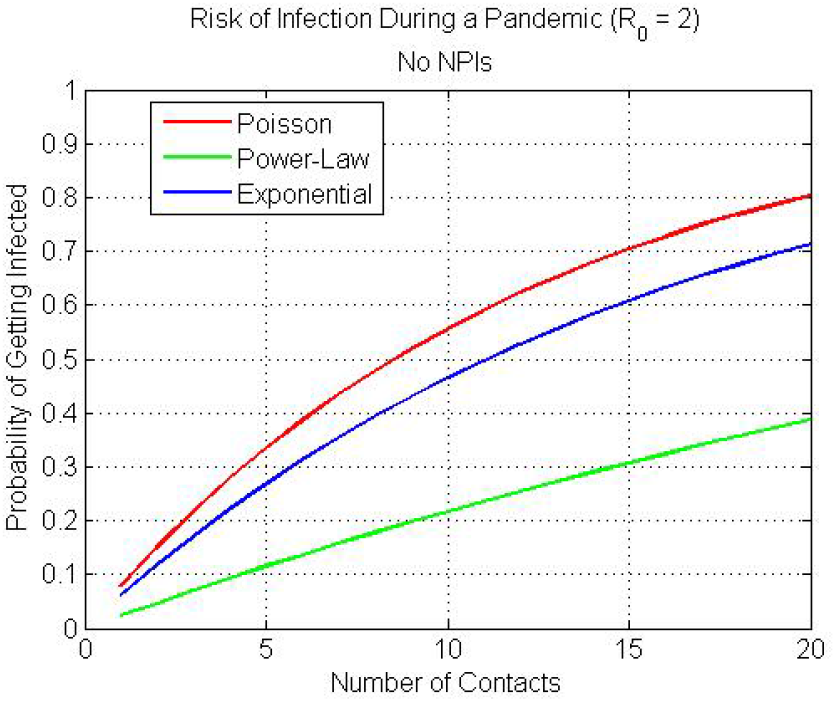
**Individual risk of an infection during a pandemic based on the number of contacts for three types of contact networks**

For uniform social distancing, it is assumed that the probability that a vertex of degree *k* is active is *b*_*k*_ = *b*, where 0 ≤ *b* ≤ 1, so 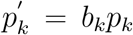. Basically, social distancing is applied to every individual regardless of the number of contacts an individual may have. Fig. 7 depicts the fraction of the population affected by a pandemic as a function of *R*_0_ for different degrees of uniform social distancing for three types of networks. Uniform social distancing has the effect of shifting the onset of a pandemic to larger effective *R*_0_ values. This is because the effective critical transmissibility threshold *T*_*c*eff_ is now greater than the baseline critical threshold *T*_*c*_ before intervention is imposed. Asymptotic results for the Poisson network are consistent with SIR/SEIR compartmental model results. In each case, the fraction of the affected population is reduced exactly by the amount of imposed social distancing. The exponential network shows similar trends although its asymptotic values are smaller than the Poisson network because the population is not well-mixed. The power-law network shows little variation in its pandemic onset or its peak (at *R*_0_ = 6) for a 20% increase in social distancing from the baseline. Even at 40% social distancing, the percent affected population is only somewhat reduced, although its pandemic onset is shifted to a larger *R*_0_ value. As explained previously, this is due to a minority of super-spreaders that add a degree of robustness to the network

**Figure 7.**
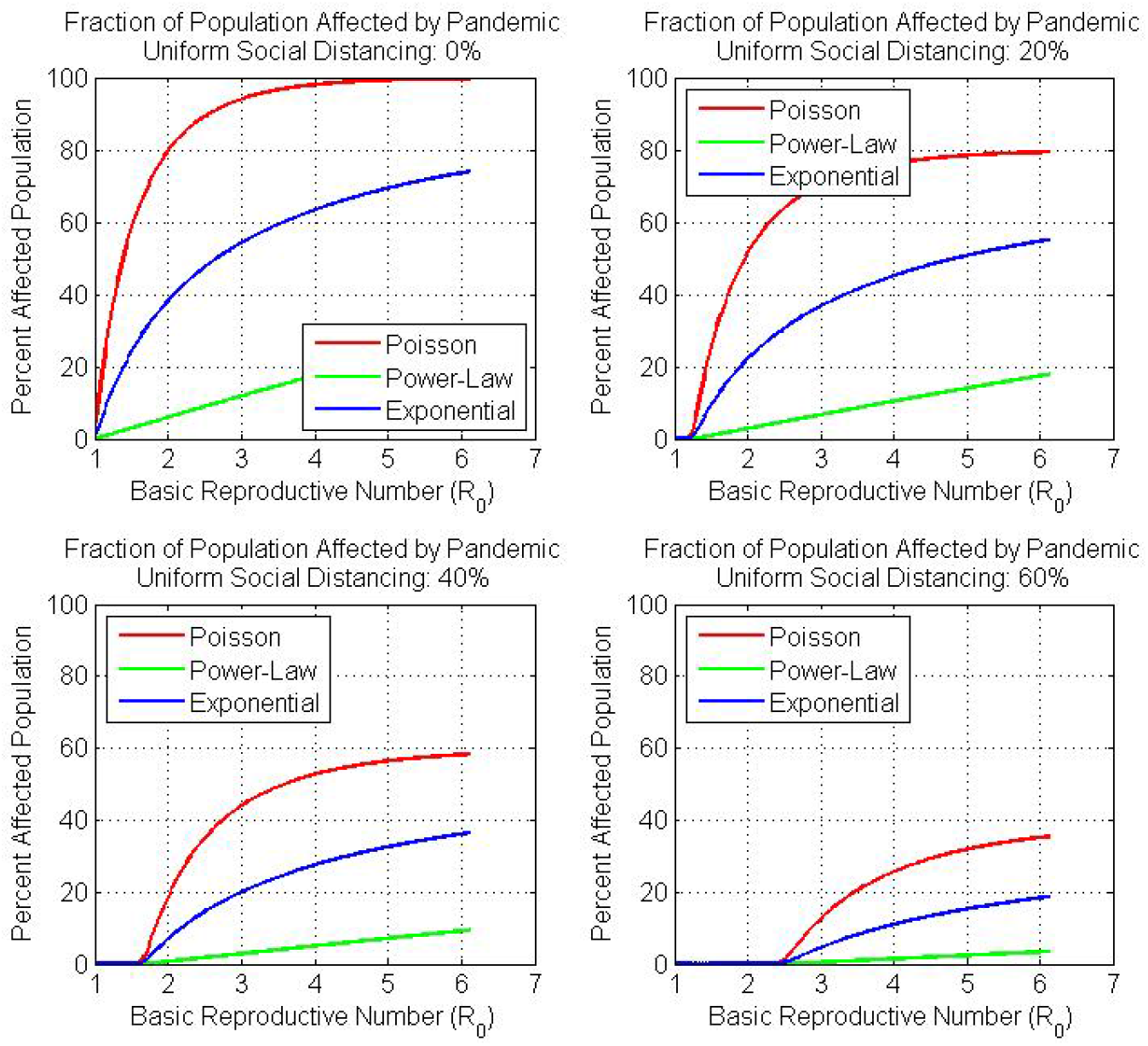
**Pandemic size as a function of** *R*_0_ **for three types of contact networks assuming uniform social distancing**

For directed social distancing, it is assumed that *b*_*k*_ = 1 for 0 ≤ *k* ≤ *K*_max_ and *b*_*k*_ = 0, otherwise. Basically, all individuals with contact degree greater than *K*_max_ are distanced. Note moved, or 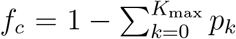. Fig. 8 illustrates the fraction of that *K*_max_ is related to the fractional number *f*_*c*_ of nodes rethe population affected by a pandemic as a function of *R*_0_ for different degrees of directed social distancing for three types of networks. Directed social distancing has a large effect on the onset of a pandemic for both the exponential and power-law contact networks. For a 10% reduction in social distancing for high social contact individuals, the affected population is reduced from 75% to 50% for an exponential network and 30% to zero for a power-law network when *R*_0_ = 6. The affected population for a Poisson network is reduced by only 10%. Although not depicted in this figure, it can be shown that for a 1.5% reduction in directed social distancing, there is a 25% reduction in the affected population for a power-law network when *R*_0_ = 6. For a 2% reduction in directed social distancing, none of the population is affected for *R*_0_ ≤ 6. However, the exponential network shows only a 5% reduction and the Poisson network shows only a 2% reduction in the affected population, respectively. The power-law network is the least robust to the removal of high contact nodes. This is an important result that could impact the way contact tracing is performed.

**Figure 8.**
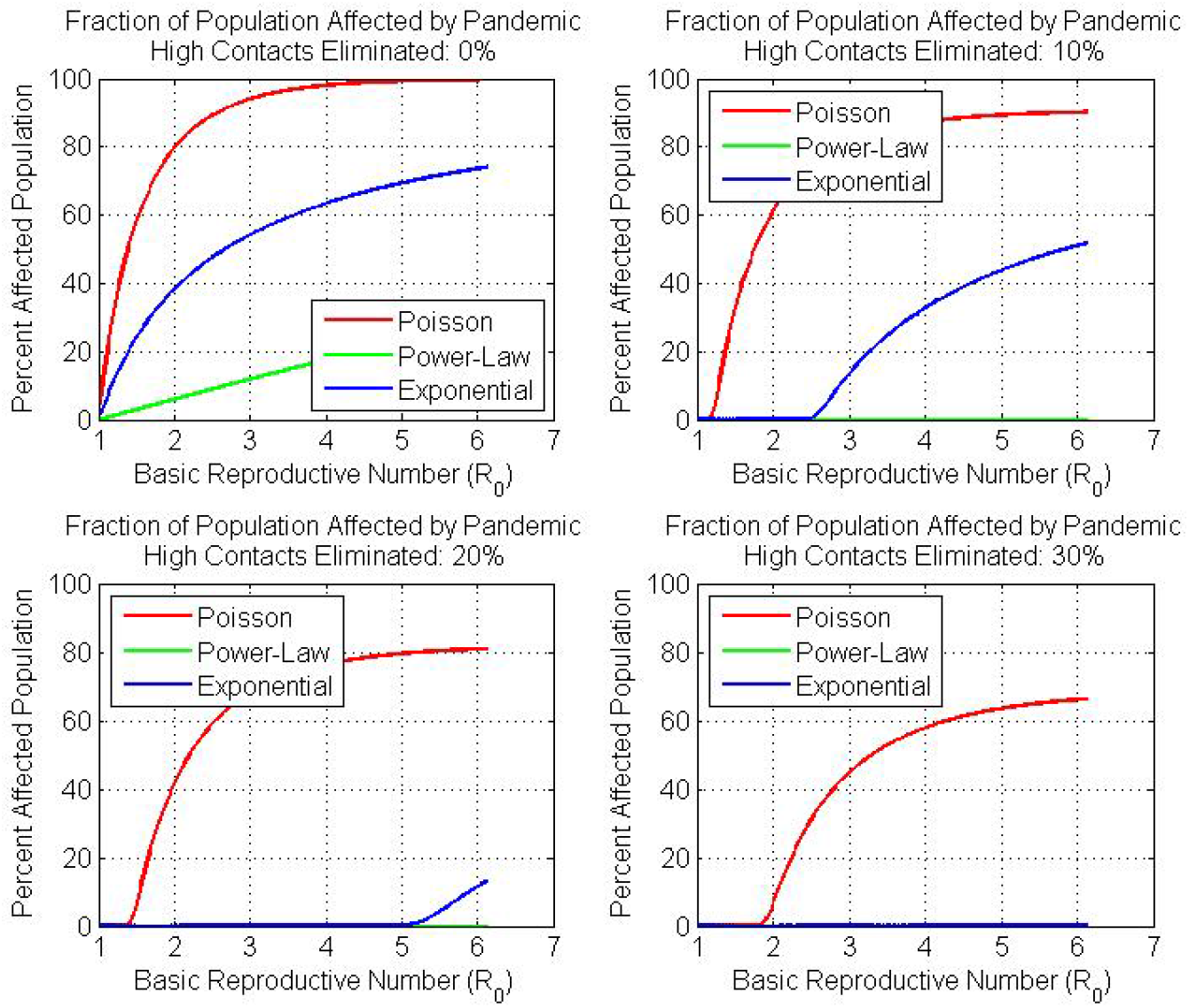
**Pandemic size as a function of** *R*_0_ **for three types of contact networks assuming directed social distancing**

### 3.2. Time-Dependent Disease Spread in Social Networks

Some illustrative results using the time-dependent disease spread model are described below. Here, it is assumed that the critical transmissibility is *T*_*c*_ = 0.049, the observed transmissibility is *T* = 0.098 (implying *R*_0_ = 2 initially), the infectious period *T*_*r*_ is 7 days, the inception and release of (uniform) social distancing starts at week 6 and ends at week 14, respectively, and testing and contact tracing begins at week 16. Additionally, 80% of individuals are social distanced, the percent isolated per week is 20%, and the percent traced per week is 30% per week.

Figures 9 and 10 depict the susceptible, infectious, and outbreak case load probabilities for a Poisson and exponential network, respectively, for the example parameters outlined above. The pandemic begins to build after week 2 and is arrested starting at week 6 as a result of social distancing. Social distancing is relaxed at week 14 with a consequent buildup in infections until testing and contact tracing are initiated at week 16. There is a large decline in the susceptible probability due to contact tracing around week 20. Since testing and contract tracing is assumed to continue over the model run (50 weeks), the outbreak case load reaches a steady state. Contrasting the differences between the Poisson and exponential networks, it is apparent that the number of outbreak cases is smaller for the exponential network and remediation strategies such as social distancing, testing, and contact tracing are more effective for the exponential network. Recall that this network does not assume a well-mixed population where all individuals are equally likely to become infected. The power-law network (not shown) has an even smaller number of outbreak cases.

**Figure 9.**
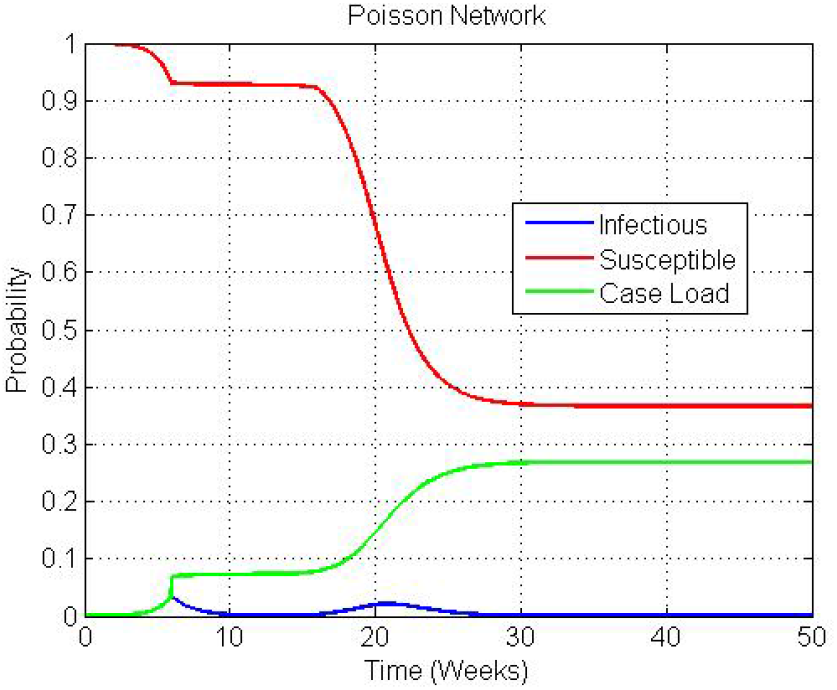
**Susceptible, infectious, and outbreak case load probabilities as a function of time for a Poisson network based on example parameters**

**Figure 10.**
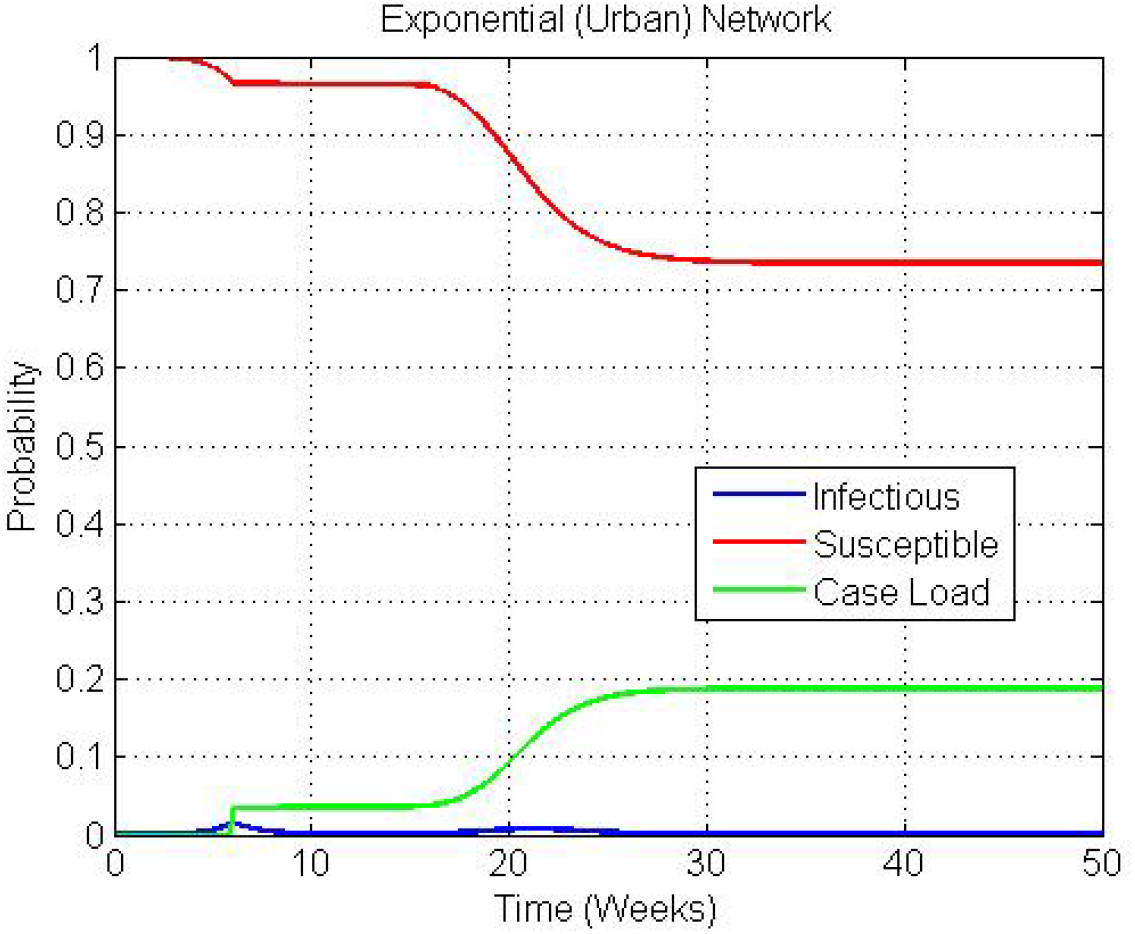
**Susceptible, infectious, and outbreak case load probabilities as a function of time for an exponential network based on example parameters**

Fig. 11 depicts the instantaneous basic reproduction number *R*_0_ for three contact networks as a function of time for the NPI, testing, and contact tracing example outlined above. The inception and lifting of social distancing protocols are clearly evident, where *R*_0_ is sizeably reduced over a period of 2 months. Note that for all three networks, 0.3 ≤ *R*_0_ ≤ 0.4 over this interval of time. When the NPI is lifted at week 14, sheltered individuals are added back into the reservoir with a subsequent increase in *R*_0_. This pool slowly attrits from infections until week 16 when testing and contact tracing is initiated. A combination of testing and contact tracing lowers *R*_0_ to below unity near week 20, at which point, the disease ceases to spread. It is interesting to note that the power-law network has consistently lower *R*_0_ values except during testing and contact tracing. This is because the rather low rates of testing and tracking miss a number of super-spreaders that are more prevalent in a power-law network.

**Figure 11.**
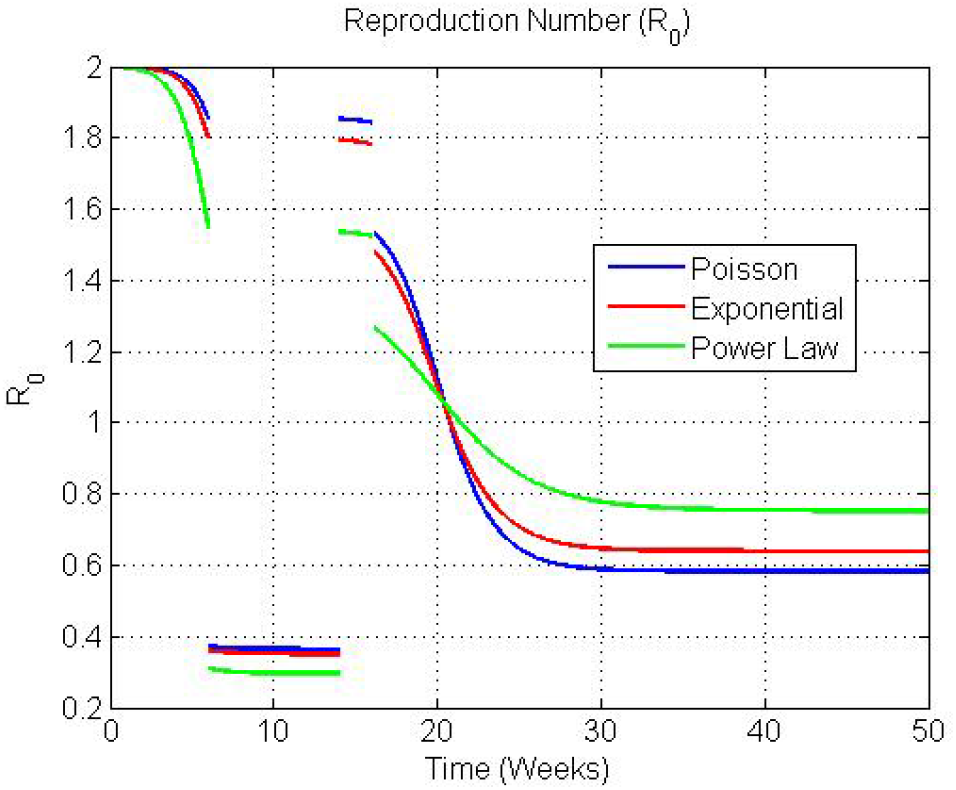
**Instantaneous basic reproduction number** *R*_0_ **as a function time for three contact networks based on example parameters**

### 3.3. Bayesian-based Estimation of Contact Transmissibility

Some illustrative results using the Bayesian-based estimation procedure for contact transmissibility are discussed below. Figures 12 and 13 illustrate particular examples of the approach for Massachusetts and New York given collected outbreak case histories from each state over a 2-3 month period starting in February 2020. It is assumed that the critical transmissibility *T*_*c*_ = 0.049 and the infection period *T*_*r*_ = 7 days. The models that best fit the data are used for each state – a Poisson network for Massachusetts and an exponential network for New York. The estimated transmissibility for Massachusetts is 0.089, which translates into a *R*_0_ value of 1.8. Similarly, the estimated transmissibility for New York is 0.185, which translates into a *R*_0_ value of 3.8. Both networks provide reasonable fits to their respective observed differential outbreak case histories Δ*C*(*t*) and reinforces the notion that the rapidity of disease spread in New York was much more severe.

**Figure 12.**
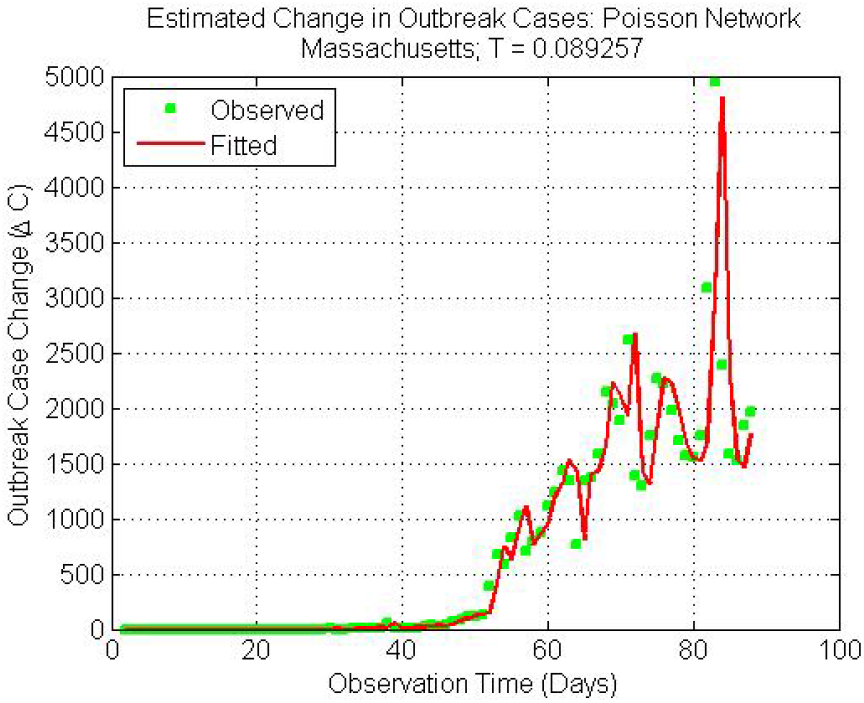
**Estimated and predicted change in outbreak cases** Δ*C*(*t*) **for a Poisson network based on Massachusetts State outbreak data**

**Figure 13.**
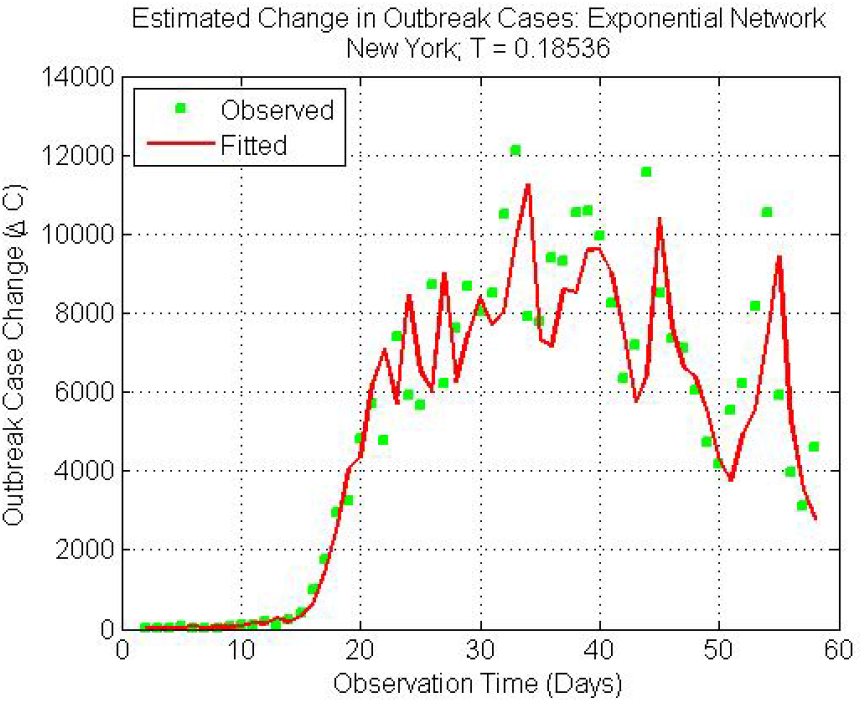
**Estimated and predicted change in outbreak cases** Δ*C*(*t*) **for an exponential network based on New York State outbreak data**

The instantaneous basic reproduction number *R*_0_(*t*) can also be estimated by tracking the differential outbreak cases over time using a particle filter. Figures 14 and 15 illustrate the estimation technique for New York given the collected outbreak history. The fit to the observed differential outbreak case data Δ*C*(*t*) is quite good. In addition, the instantaneous *R*_0_(*t*) values are consistent with the average *R*_0_ value using an exponential network model depicted in Fig. 13. However, Fig. 15 is more illustrative because it allows one to examine the trend in *R*_0_ over the progression of the pandemic. In this case, there is a downward trend after reaching a peak of approximately 3.5. There is also an up tick in *R*_0_ at later times probability due to an increase in testing. Note that estimation errors may result in negative *R*(*t*) values that are not realistic.

**Figure 14.**
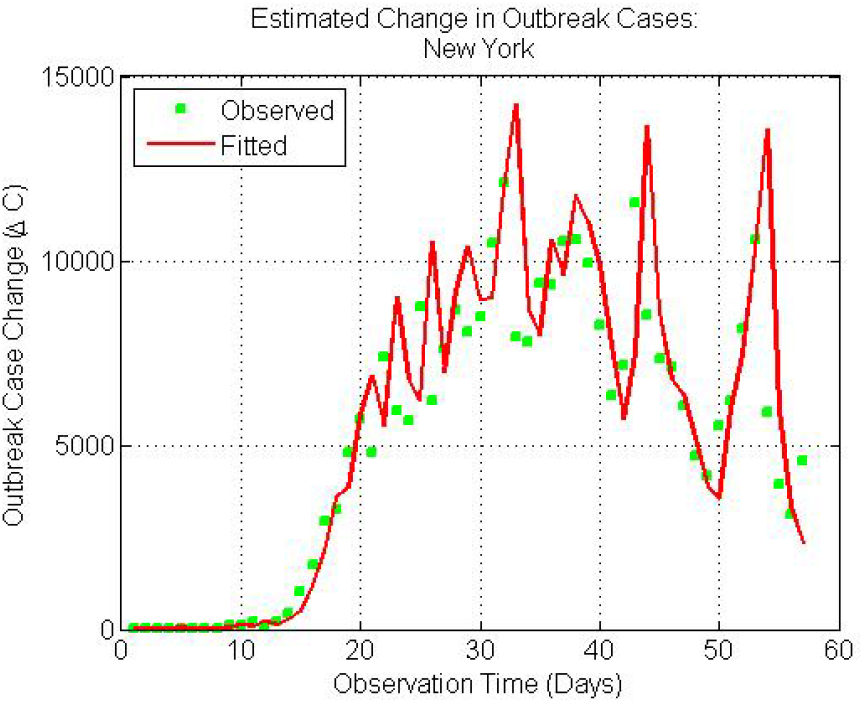
**Estimated and predicted change in outbreak cases** Δ*C*(*t*) **based on New York State outbreak data using a particle filter**

**Figure 15.**
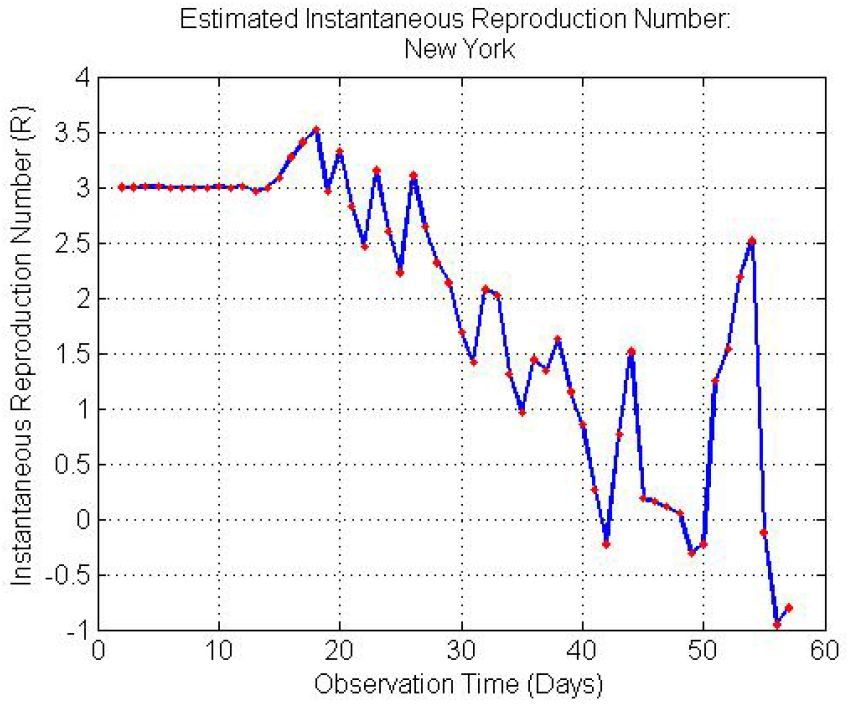
**Estimated instantaneous basic reproduction number *R*_0_ based on New York State outbreak data using a particle filter**

## 4. Conclusion

In the present work, we developed and illustrated a coarsegrain analytic modeling procedure that can be used to assess alternative strategies of implementing and subsequently lifting non-pharmaceutical interventions in response to the COVID-19 pandemic. This work was based on developing a multidimensional view of the problem by examining steady-state and dynamic disease spread using a network-based approach. The steady-state models, based on percolation theory, highlighted the previously known result that social contact structure is a key factor in the size of an outbreak. In addition, it was shown that the social contact structure influences the types of social distancing protocols that are deemed most effective. The dynamic models, based on a stochastic reformulation of the SIR equations, further extended the work to include the effects of lifting non-pharmaceutical interventions and the importance of testing and contact tracing in reducing the overall infection rate. Providing a realistic assessment of the basic reproduction number *R*_0_ is also important in gauging the severity of the outbreak within a specific geographic area. A Bayesian-based estimation procedure was developed to estimate both the average and instantaneous basic reproduction number from outbreak case histories at a state and county-wide level. These estimates can be used to seed other models or analysis procedures.

No single model is a panacea. Therefore, we advocate an ensemble modeling approach based on a combination of analytic and fine-grain agent-based models (ABMs). As outlined in a companion paper, this approach has the potential to provide valuable insights into disease spread and the effectiveness of nonpharmaceutical interventions. It could prove to be a valuable tool for decision-makers in conjunction with empirical analysis.

## Data Availability

The validation data is publicly available and cited in the paper.

